# Correlates of obstetric violence by maternity care providers during antenatal consultations and childbirth in two referral hospitals in Bafoussam (West-Cameroon)

**DOI:** 10.64898/2026.01.23.26344699

**Authors:** Alima Jean Marie, Ikei Solange Azenoi, Fouogue Tsuala Jovanny, Tamo Fogue Yannick, Fouedjio Jeanne Hortence, Kenfack Bruno

**Affiliations:** Faculty of Medicine and Pharmaceutical sciences, University of Dschang; Department of Gynecology and Obstetrics, Bafoussam Regional Hospital; Department of Psychiatry, Bafoussam Regional Hospital; Faculty of Science of Education, University of Yaoundé 1

**Author notes:** Corresponding author: Alima Jean Marie, Faculty of Medicine and Pharmaceutical Sciences, University of Dschang-Cameroon.

**Keywords:** Obstetric violence, pregnancy, violence, abuse, disrespect and abuse, mistreatment, Bafoussam

## Abstract

**Objective:** determine the prevalence of Obstetric Violence (OV) and its correlates in Bafoussam.

**Methodology:** we conducted a quantitative hospital-based survey at the Bafoussam Regional Hospital (BRH) and the Mifi District Hospital (MDH) from October 15th to December 15th 2023.Our sampling was consecutive and exhaustive including women in post-partum wards. We conducted face-to-face interviews using a pretested questionnaire and completed data using medical records. We performed chi square test of independence and, binary and multiple logistic regressions to measure associations between variables using SPSS 26.0 software (IBM, New York, NY). A p-value < 0.05 was considered significant.

**Results:** Five hundred and twelve women were included. The proportion of women who experienced OV during antenatal care (ANC) was 8.1% (40/492). Of those women, 16 (40.0%) continued their ANC in another health facility. The proportion of intrapartum OV was 54.7% (274/512). The most frequent forms of OV during this period were: perineal suture without anesthesia (43.6%), no greetings nor self-introduction by the personnel (38.2%), digital vaginal examination without consent (24.6%) and the non-preservation of intimacy (21.8%). Inability to pay for healthcare services was associated to both antenatal OV (aOR: 2.9(1.3 - 64), p: 0.01) and intrapartum OV (aOR: 19.6 (95 %(1.7-26.9), p: 0.01).

**Conclusion:** OV is quite common and varied in Bafoussam. Its drivers and impact should be investigated to inform effective mitigation strategies.

**Synopsis:** This study found that half of women suffer OV during childbirth and one in ten women during ANC. Poor women were disproportionately affected.

## Introduction

Obstetric violence(OV) is any violence against women perpetrated by obstetric care providers regarding the body and reproductive processes of the woman.(1) Despite being unacceptable to health care providers (HCP) and even the society, OV still exists in various forms worldwide.(2)

In 2015, the International Federation of Gynecology and Obstetrics (FIGO), together with several other international bodies, proposed the Mother and Baby Friendly Birth Facility (MBFBF)based on seven categories identified as: disrespect and abuse (Physical abuse; Non-consented care; Non-confidential care; Non-dignified care; Discrimination based on specific patient attributes; Abandonment of care and Detention in facilities(3) (4). OV has been identified as a barrier among others to accessing health facilities thereby increasing maternal morbi-mortality which remains a burden in Cameroon.(5) This hinders the attainment of target 3.1 of sustainable development goal 3 which is focused on reducing maternal mortality by reducing all forms of violence against women (6) (7) (8). Providing respectful maternity care is vital for promoting timely care-seeking behavior, thus ensuring the health and well-being of (woman-baby dyads) (9). OV is a global phenomenon with prevalence ranging from 18.3% (in Brazil) (10) to 75.1% (in Ethiopia) (11).

Obstetric violence has been an under-looked obstacle to quality maternal health care services. There is limited evidence on OV in Cameroon and particularly, in the West Region. Hence, this pioneer study, aiming at assessing the prevalence of OV and its correlates in 2 hospitals in the West Region of Cameroon.

## Materials and Methods

### Study design

This was a quantitative cross-sectional hospital-based study.

### Study setting

The study was conducted in 2 hospitals of the capital city of the West Region of Cameroon; the Bafoussam Regional Hospital (BRH) and the Mifi District Hospital (MDH).These are 3^rd^ and 4^th^ category hospitals and serve as referral hospitals at different levels for the Region with about 1200 and 1560 births per year respectively.

### Study period

Data was collected from October 15th to December15th 2023.

#### Study population

##### Inclusion criteria

all consenting mothers who gave birth vaginally or through caesarean section in the study sites during our study period were eligible.

##### Sample size

A single population proportion formula was used to estimate the sample size. We assumed a prevalence of OV of 50% given that we found no prior study on the topic in Cameroon. We considered a 5% error margin, a 95% confidence interval and a 10% non-response rate for the calculations. We were expected to recruit at least 424 participants in both hospitals.

##### Sampling technique

Participants were included consecutively until our required sample size was reached.

### Methods and procedures

Data was collected by 7 trained (on violence and ethics) data collectors who had no prior contact with the participants during ANC and childbirth. Our questionnaire was adapted from an original version validated in Brazil for the WHO Multi-country Study on women’s Health and Domestic Violence against Women (7) that addressed psychological, physical and sexual violence perpetrated against women in different social contexts. This standardized structured questionnaire was reviewed, translated to French and back translated to English and pretested on 50 participants who were not included in the final sample. All these to ensure accuracy, cognitive understanding, and cultural acceptability. Every day the data collectors identified soon to be discharged women whom they invited to participate. Consenting women were interviewed in a private area of the hospital. All filled questionnaires were verified and validated by the Principal investigator (PI) daily. The data was entered into a predesigned data entry sheet then matched and crosschecked by the PI for coherence.

### Statistical analysis

Data was analyzed using SPSS 26.0 (IBM, New York, NY). Proportions were presented for categorical variables while means were presented for continuous variables. With a statistically significant threshold set at p-value< 0.05, the chi square test of independence, binary and multiple logistic regressions and their odds ratio with their 95% CI were reported.

### Ethical consideration

Ethical clearance was obtained from the West Region Ethical Committee (N°/921/30/08/2023/CE/CRERSH-OU/VP).Official authorizations were obtained from the directors of both sites. Confidentiality and anonymity were ensured by conducting interviews in a private area with the respondent alone. The questionnaires were coded. The information notice was clearly explained to every woman and time was given them to freely consent or not.

### Participants’ benefit

All participants were given information on available services where there could get help if the need arose. For victims of OV the attending physician was notified in order to determine the need for monitoring, refering or a consultation liaison with the psychology service of the BRH where follow-up support was provided as need arose.

## Results

### Characteristics of the study population

**Figure 1:**
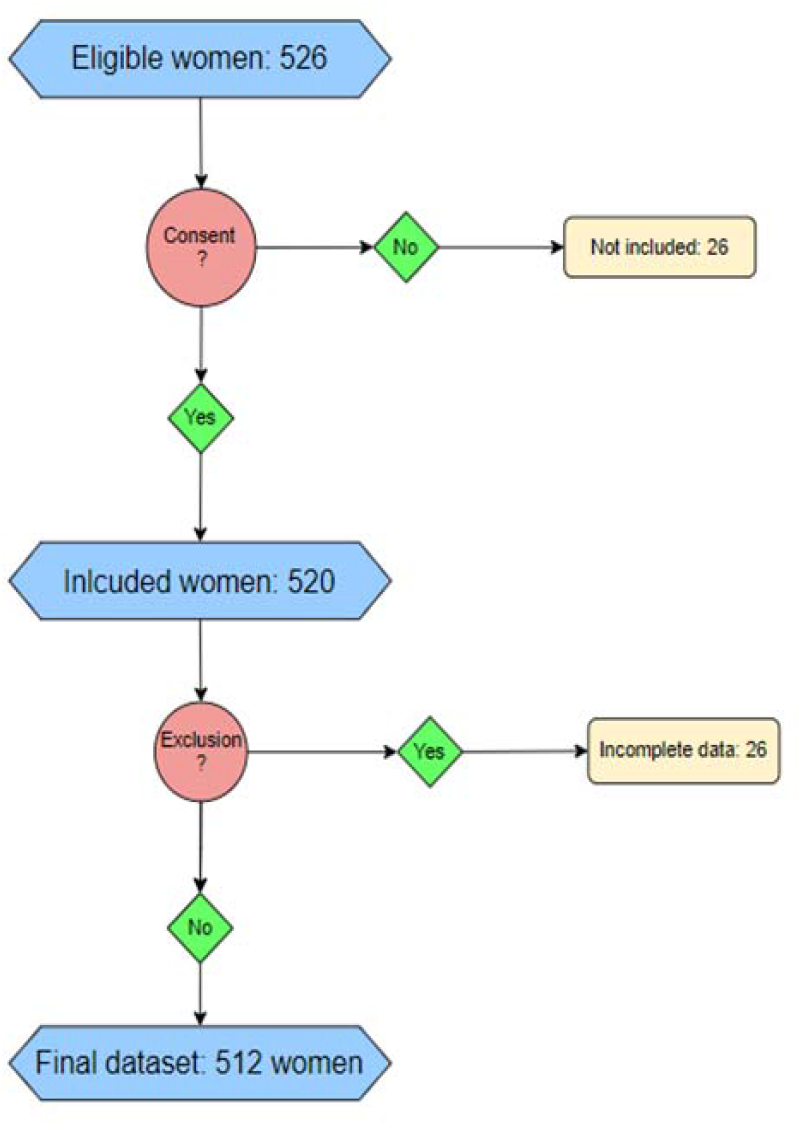
the inclusion flow diagram.

Median (interquartile range) was 27 (23 - 31) years and 148 (28.9%) were between 21 and 25 years of age. Majority of the participants, 451(87.7%) were from West Region of Cameroon. Regarding the marital and employment status, 349 (68.2%) were in a union and 147 (28.7%) were housewives. Majority of our participants 496 (96.9%) were Christians. As concerns the educational status, 486 (94.9%) had attained secondary education. Sixty percent (307) declared that they were not able to pay their hospital bills.

The average gestity of our participants was 2.8 pregnancies and the mean parity was 2.4 births. Fifteen (2.9%) had not had antenatal care while 353 (71.0%) had at least 5 ANC visits. Of those who had ANC visits, 320 (64.4%) were attended to by midwives.

During labor, 384 (75.0%) were followed up by a female personnel and 359 (70.1%) were assisted in childbirth by a female personnel. Also, 391(76.4%) were attended to during childbirth by midwives. Four hundred and forty-two mothers spent between 1and 12 hours in the hospital before childbirth and in 401 (78.3%) had a vaginal delivery. All these are presented in table 1.

**Table 1:** Characteristics of the study population (N=512)

| Variables (missing values) | Categories | Frequencies n (%) |
| --- | --- | --- |
| Socio demographic (N = 512) |  |  |
| Age (0) | 15 - 20 | 74 (14.5) |
|  | 21-25 | 148(28.9) |
|  | 26-30 | 142(27.7) |
|  | 31 - 35 | 90(17.6) |
|  | 36-40 | 45(8.8) |
|  | 41 - 45 | 11(2.1) |
|  | ≥ 46 | 2(0.4) |
| Region of origin (1) | West | 451(87.7) |
|  | Other regions | 60(12.1) |
|  | Foreigner | 1(0.2) |
| Marital status(0) | In a union | 349(68.2) |
|  | Single | 160(31.2) |
|  | Widowed | 2(0.4) |
|  | Divorced | 1(0.2) |
| Occupation(0) | Housewife | 147(28.7) |
|  | Employed | 100(19.5) |
|  | Self-employed | 139(27.2) |
|  | Student | 101(19.7) |
|  | Others | 25(4.9) |
| Religion(0) | Christian | 496(96.9) |
|  | Muslim | 14(2.7) |
|  | Others | 2(0.4) |
| Educational background(0) | No formal education | 5(1.0) |
|  | Primary Education | 21(4.1) |
|  | At least secondary | 491 (94.9) |
| Ability to pay for care (0) | Yes | 105(40.0) |
|  | no | 307 (60.0) |
| ANC<sup>a</sup> | Yes | 497(97.1) |
|  | No | 15(2.9) |
| **ANC (497)** |  |  |
| Number of ANC | 1-4 | 144(29.0) |
|  | ≥ 5 | 353 (71.0) |
| Qualification of service provider during ANC | Nurses | 87(17.5) |
|  | Midwife/male midwife | 320(64.4) |
|  | General practitioner | 26(5.2) |
|  | Obstetrician | 63(12.7) |
|  | Nursing attendant | 1(0.2) |
| Gender of the service provider during ANC | Male | 89(17.9) |
|  | female | 381(76.7) |
|  | Both | 27(5.4) |
| **Labor and childbirth (N=512)** |  |  |
| Gender of provider in labor room | Male | 125(24.4) |
|  | Female | 359(70.1) |
|  | both | 28 (5.5) |
| Time before delivery (hours) | 1H – 12 H | 442 (86.3) |
|  | 13H – 23H | 41 (8.0) |
|  | 1 – 2 days | 26 (5.1) |
|  | ≥ 3 days | 3(0.6) |
| Period of delivery | Daytime (6H – 18H) | 297 (58.0) |
|  | Night (19H – 5H) | 215(42.0) |
| Day of delivery | Business day | 390(76.2) |
|  | Weekend/holidays | 122(23.8) |
| Qualification of provider during childbirth | Nursing assistant | 3(0.6) |
|  | midwife | 391 (76.4) |
|  | Obstetrician | 58(11.3) |
|  | Nurse | 12(2.3) |
|  | General practitioner | 48(9.4) |
| Method of deliver | Vaginal delivery | 401 (78.30) |
|  | Assisted delivery | 6(1.2) |
|  | Cesarean section | 105(20.5) |

<sup>a</sup>ANC: antenatal care

### Prevalence and features of OV

Of the 512 participants 8.1% reported OV during ANC and 54.7% during childbirth.

### Obstetric violence during antenatal care

Forty participants reported OV during ANC. amongst them 17(42.5%) were scolded followed by 15(37.5%) who were poorly received and 5 (12.5%) who felt neglected in their care as depicted in table 2.

**Table 2:**
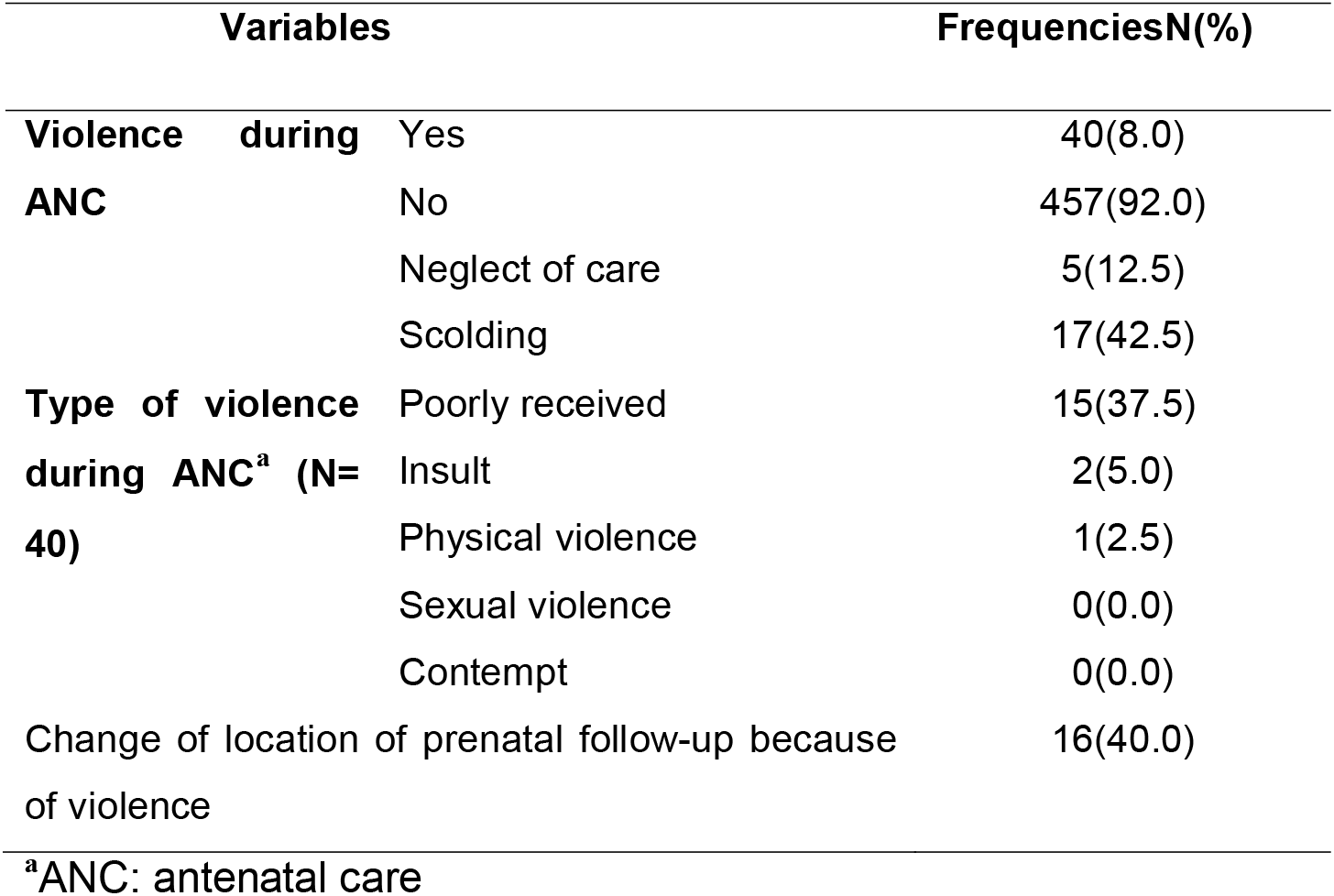
Distribution of participants by type of OV during ANC in Bafoussam.

### Prevalence and various forms of OV

Physical abuse was the most frequent category of OV followed by non-consenting care and impoliteness, then non confidential care, sexual violence and in fifth position, non-dignified care as depicted in Table 3.

**Table 3:** Categories and subtypes of obstetric violence(N=280)

| Categories and subtypes | Frequencies n (%) |
| --- | --- |
| **Physical abuse** |  |
| Yes | 159(56.8) |
| No | 121(43.2) |
| - physically hit or slapped you | 5(1.8) |
| - Separated mother from baby without any medical indications | 1(0.4) |
| - No administration of pain relief treatment | 4(1.4) |
| - Denied from food or fluids in labor without medical indication | 38(13.6) |
| - Suturing without anesthesia | 122(43.6) |
| - Episiotomy without prior information | 11(3.9) |
| **Non dignified care** |  |
| yes | 37(13.2) |
| no | 243(86.8) |
| - HCP<sup>a</sup> shouted at or scolded you | 32(11.4) |
| - HCP made negative comments about you | 2(0.8) |
| - Support staff insulted you and your companion | 1(0.4) |
| - no demonstrating caring in culturally appropriate ways | 2(0.8) |
| **Non confidential care** |  |
| Yes | 82(29.3) |
| No | 198(70.7) |
| - HCP did not use drapes or coverings to protect mother's privacy (non-respect of intimacy) | 20(7.1) |
| - HCP discussed your private health information in a way that others could hear. | 6(2.1) |
| - your ward could not allow the respect of intimacy | 61(21.8) |
| **Non consenting care and impoliteness** |  |
| Yes | 126(45.0) |
| No | 154(55.0) |
| - HCP did not introduce themselves nor greeted the mother and her carers | 107(38.2) |
| - HCP did not encourage mother to ask questions | 8(2.9) |
| - HCP did not respond to mother's question with politeness | 4(1.4) |
| - HCP did not explain what is being done and what to expect throughout labor and birth | 9(2.9) |
| - no information given nor seeking consent | 8(2.9) |
| - HCP did not permit mother to choose her birth position | 9(3.2) |
| **Abandonment of care** |  |
| Yes | 30(10.7) |
| No | 250(89.3) |
| - HCP ignored you when you called for help | 17(6.1) |
| - left unattended during the second phase of labor | 11(3.9) |
| - HCP did not administer my treatment | 5(1.8) |
| **Sexual violence** |  |
| Yes | 73(26.1) |
| No | 207(73.9) |
| - HCP frequently performed vaginal examinations without my consent | 69(24.6) |
| - HCP touched me while I have been refusing | 4(1.4) |
| - HCP made negative comments on my intimacy | 1(0.4) |
| **Discrimination** |  |
| Yes | 1(0.4) |
| No | 279(99.6) |
| **Detention in facility** |  |
| Yes | 15(5.4) |
| No | 265(94.6) |

<sup>a</sup>HCP: Health care provider

### Correlates of obstetric violence

Of all the factors associated to OV from the bivariate analysis on “Table 1S” and “Table 2S”, only the inability to pay for services stood out as an associated factor after the multivariable logistic regression as seen on table 4.

**Table 4:** Multivariate regression analysis of factors associated to obstetrical violence during ANC, labor and delivery.

| Exposure variables | OV<sup>a</sup> during labor and childbirth |  | Adjusted OR<br>(95%CI) | Adjusted P<br>value |
| --- | --- | --- | --- | --- |
|  | Yes | No |  |  |
| Inability to pay for care | 195 (69.6) | 112 (48.3) | 19.6 (1.7-26.9) | 0.010 |
| Age ≤ 30 | 214 (76.4) | 150 (54.7) | 1.1(0.7– 1.9) | 0.541 |
| Number of ANC:1- 4 | 89 (33.1) | 55 (24.1) | 1.3 (0.9 – 2.1) | 0.138 |
| Same provider at labor & childbirth | 133 (47.5) | 135 (58.2) | 0.8 (0.5 – 1.2) | 0.398 |
| Parity: 1-2 | 193 (68.9) | 120 (51.7) | 1.4 (0.9 – 2.2) | 0.121 |
| Time before delivery(13-23h) | 29 (10.8) | 12 (5.6) | 1.9 (0.95 – 4.06) | 0.069 |
|  | OV during ANC |  |  |  |
|  | Yes | No |  |  |
| Inability to pay for care | 32 (80) | 263 (57.5) | 2.9 (1.3-64) | 0.01 |

<sup>a</sup>OV: Obstetric violence

## Discussion

This study revealed that the prevalence of OV was 8.1% during ANC. The commonest forms of OV during ANCs were: shouting (42.5%), poor reception (37.5 %), and neglect (12.5%). Forty percent of women who experienced OV continued their ANC in another facility. More than half of women experienced OV during childbirth and the commonest were: perineal repair without anesthesia (43.6%), no greetings nor self-introduction by providers (38.2%), unconsented digital vaginal examination (24.6%) and non-preservation of intimacy (21.8%). The inability to pay for health care services was associated with OV.

The participants were interviewed postpartum. This could be the reason for the low prevalence of VOs during prenatal consultations, probably due to women forgetting the events that marked their prenatal follow-up 9 months earlier.

Incidentally, the high prevalence of OV during childbirth could be attributed to the stress of childbirth on both the mother and the HCP. This high prevalence in our study is close to what has been reported in African countries with similar socio-cultural features and reproductive health indicators; Kenya, Nigeria, Malawi and Ethiopia. (12) (13) (14) (15)

Physical violence was experienced by more than half of the victims of OV and among these; perineal tear repair without anesthesia was the most represented. Our results were similar to those of Wasihum et al in Ethiopia (12) but higher than those of other studies carried out in Nigeria (13) and Tanzania (16). This questionnable practice could be motivated by the need to avoid the extracost of the required materials and drugs for the procedure.

Some were denied food or fluid during labor without a medical indication.This rate is lower than was reported in a study in Jordania (17), Nigeria (13) and south Ethiopia but higher than was reported in a study in rural Kenya (14). This practice, inherited from old recommendations to avoid Mendelson syndrome, is still present in our delivery rooms, and midwives seem to be slow to implement the new WHO guidelines (18).

Non-consented care was the second most represented category of OV in our study and concerned close to half of the victims. This could be underpinned by medical paternalism towards female patients. According to our findings, the next most common category of OV experienced by women was non-confidential care. This finding was higher than was reported in studies conducted in Kenya, Nigeria and Ethiopia (12) (13) (14). In our study, about one fifth of mothers complained that the delivery rooms did not permit their privacy and intimacy to be respected. This pertains to the structural conditions of the delivery room. This can also be called institutional violence which is any action committed by or in an institution or every action omitted, which leads to unnecessary physical or psychological pain (19). There is a need to rethink the physical organization of maternities and delivery rooms in order to guarantee more intimacy for parturient in our setting.

As concerns the factors associated to OV, participants who could not afford maternity care services were three times and 20 times more likely to undergo OV during ANC and delivery, respectively. This indicates the possible higher vulnerability of low-income categories to OV in Bafoussam.These results can be explained by the poor working conditions of HCP dating back to the austerity policies imposed by the Cameroon government in the 1990s (18); and as a result, an impoverished and demotivated staff (20) (21). These workers will therefore have the tendency to favor some patients compared to others due to purely economic reasons (bonuses, unofficial payment, bribing etc). Also, the low economic power of certain population is sometimes accompanied by low level of education and information as concerns their rights and their health. These factors of precarity increases the vulnerability of mothers during childbirth and thus contributing legitimately to acts of physical and verbal violence that they can suffer from health personnel.

### Strength of the study

this study is the first of its kind in our country.

### Limit of the study

recall bias: Given the long time laps between participants ANC and the interview in the early post-partum, they could have forgotten episodes of violence. The effect of that recall bias on the proportion of OV during childbirth was less important.

## Data Availability

All data produced in the present study are available upon reasonable request to the authors

## Conflict interests

The authors declare that they have no competing interests.

## Authors’ contribution

ALIMA J.M., IKEI S., FOUOGUE T.J. designed the study, coordinated the data collection and management, analyzed the data and prepared the manuscript. TAMO F. Y., FOUEDJO J.H. and KENFACK B. participated in the design of the study, data collection and revised the manuscript. All authors read and approved the final manuscript.

## Funding

This research did not receive any specific grant from funding agencies in the public, commercial or not-for-profit sectors.

## Acknowledgments

The authors are thankful to all the participants and nvestigators.

## Notes

### Competing Interest Statement

The authors have declared no competing interest.

### Funding Statement

This study did not receive any funding

### Author Declarations

Ethics committee for human health research for the west Region of Cameroon gave ethical approval for this work (N /921/30/08/2023/CE/CRERSH-OU/VP).

